# Differential association of body mass index with hypoglossal nerve stimulation efficacy by pharyngeal collapse pattern in obstructive sleep apnea

**DOI:** 10.64898/2026.01.25.26344734

**Authors:** Daniel Vena, Eric J. Kezirian, Andrew Wellman, David Kent, Mark D’Agostino, Joao L.G.C. Monteiro, A Azarbarzin, Tom Chen, Ludovico Messineo, Neda Esmaeili, Scott A. Sands, Phillip Huyett

## Abstract

**Background:** Hypoglossal nerve stimulation (HGNS) is an established surgical therapy for obstructive sleep apnea (OSA), yet treatment response is variable and appears to be influenced by both body mass index (BMI) and pattern of upper airway collapse. Whether excess body weight differentially affects HGNS efficacy across pharyngeal collapse patterns remains unclear.

**Methods:** We combined data from two independent HGNS cohorts (n=760) to examine whether the association between BMI and HGNS efficacy depends on pharyngeal collapse pattern identified by drug-induced sleep endoscopy. Collapse was categorized as predominantly laterally directed or anteroposterior (AP). Multivariable regression models assessed HGNS efficacy—quantified primarily as percent reduction in apnea–hypopnea index (AHI) on titration polysomnography and secondarily as treatment success (≥50% AHI reduction to <15 events/h)—while testing for effect modification by collapse pattern and adjusting for baseline AHI, partial collapse, surgical center, follow-up sleep study type, and prior or concomitant pharyngeal surgery.

**Results:** Increasing BMI was associated with substantially lower HGNS efficacy among patients with laterally directed collapse, whereas the relationship between BMI and efficacy was attenuated in those with AP collapse. Specifically, each 5 kg/m^2^ increase in BMI was associated with a −19.7% (95% CI: −33.2 to −6.2) greater reduction in efficacy in lateral collapse compared with −3.8% (−8.0 to 0.36) in AP collapse (interaction p=0.027). Higher BMI also corresponded to reduced odds of treatment success in lateral collapse (odds ratio 4.4 [95% CI: 1.4–14.3] per 5 kg/m^2^), with no significant association observed in AP collapse (1.1 [0.75–1.5]; interaction p=0.023).

**Conclusions:** The influence of BMI on HGNS treatment response differs meaningfully by pharyngeal collapse phenotype. Incorporating collapse pattern into BMI-based eligibility criteria may improve patient selection and optimize HGNS outcomes.

## INTRODUCTION

Unilateral hypoglossal nerve stimulation (HGNS; Inspire Medical Systems) has become an increasingly utilized therapy for obstructive sleep apnea (OSA) in patients who are unable to tolerate positive airway pressure^1^. Despite its growing use, HGNS demonstrates variable efficacy, with a substantial proportion of patients failing to achieve an adequate reduction in apnea–hypopnea index (AHI)^2–9^.

Two factors that have been consistently associated with reduced HGNS efficacy are elevated body mass index (BMI)^10^ and pharyngeal collapse patterns with a prominent lateral component, including complete concentric collapse (CCC) at the palate and complete oropharyngeal lateral wall (OLW) collapse^5,11–14^. Higher BMI is associated with increased upper airway collapsibility, which may reduce the ability of HGNS prevent upper airway collapse^15–18^. Similarly, lateral pharyngeal collapse may limit HGNS effectiveness, as stimulation primarily produces anteroposterior displacement of the tongue and adjacent structures, potentially leaving laterally directed collapse unresolved. However, the interaction between BMI and pharyngeal collapse pattern in determining HGNS efficacy has not been well characterized.

Clarifying this interaction has important implications for clinical decision-making. Patients with higher BMI may still respond favorably to HGNS when collapse occurs at sites that are more directly targeted by stimulation, such as the tongue base. Conversely, patients with laterally directed collapse may experience good treatment response in the setting of lower BMI. Although lateral collapse patterns are more common at higher BMI, obesity does not uniformly predict the presence of lateral collapse, underscoring the need to consider both factors together when evaluating HGNS candidacy^19^. This issue is particularly relevant given recent changes to regulatory guidance that have expanded BMI eligibility for HGNS despite limited efficacy data.

The present study evaluates the relationship between BMI and HGNS efficacy across different pharyngeal collapse patterns using pooled data from two independent HGNS cohorts. We hypothesized that greater BMI would be associated with reduced HGNS efficacy in collapse patterns that are less responsive to stimulation, such as CCC and complete OLW collapse, but would have a smaller effect in predominantly anteroposterior collapse patterns.

## METHODS

### Study Design and Cohorts

This study pooled data from two independent cohorts of patients treated with HGNS for OSA: a multicenter retrospective cohort (n = 343)^5^ and a prospective, single-center observational cohort (n = 419) drawn from the Mass Eye and Ear Sleep Surgery Registry. Both cohorts included patient demographics, BMI, pharyngeal collapse characteristics assessed by drug-induced sleep endoscopy (DISE), and baseline and post-treatment measures of OSA severity. BMI data were missing for two patients in the retrospective cohort, resulting in a total analytic sample of 760 patients.

In the prospective cohort, 560 patients who underwent HGNS implantation between October 2019 and June 2024 were enrolled in a study evaluating factors associated with HGNS efficacy (IRB: 2020P000156). Of these, 419 patients had completed post-implantation sleep testing and were included in the present analysis. Patients who did not undergo follow-up sleep testing due to patient preference or insurance denial remained under routine clinical follow-up. Patients who underwent tonsillectomy (n = 35), functional expansion pharyngoplasty (n = 10), or partial uvulectomy (n = 3) either prior to or concomitant with HGNS implantation without repeat DISE were included, with adjustment for these procedures in statistical analyses as described below.

### HGNS Implantation and Sleep Study Assessment

All patients underwent DISE prior to HGNS implantation. Baseline AHI was assessed using either a clinical home sleep apnea test or an in-laboratory polysomnogram (PSG) performed prior to DISE evaluation. Patients subsequently underwent unilateral HGNS implantation using a standard surgical approach (Inspire Medical Systems, Golden Valley, MN). Device activation occurred approximately one month after implantation, followed by post-activation sleep testing to assess and optimize therapeutic efficacy.

Most patients underwent an in-laboratory titration PSG (tPSG) after device activation. A subset of patients in the prospective cohort (n = 62) elected to undergo home sleep apnea testing following subjective optimization of therapy at home based on symptom improvement and tolerability. Across cohorts, post-activation care pathways varied over time, with tPSG initially recommended at one month after activation and later updated to approximately three months. For patients undergoing tPSG, AHI was calculated during periods of optimal stimulation.

In the prospective cohort, sleep studies were scored according to American Academy of Sleep Medicine guidelines, with hypopneas defined by a 3% oxygen desaturation criterion^20^. In the multicenter retrospective cohort, sleep study scoring was not standardized across sites; however, scoring definitions for follow-up studies were consistent with those used at baseline within each patient.

### Assessment of Pharyngeal Collapse

Pharyngeal collapse patterns were assessed using DISE and classified according to the VOTE system^21^. In the prospective cohort, DISE was performed using standardized techniques, and VOTE classifications were assigned at the time of the procedure by the treating surgeon. In the multicenter cohort, DISE examinations were performed according to standard practice at each participating center, and consensus VOTE scores were generated by four physician reviewers who were blinded to treatment outcomes^5^.

### Categorization of Collapse Patterns

For analytic purposes, collapse patterns were grouped into two primary categories: lateral collapse and AP collapse. Lateral collapse included CCC and complete OLW collapse. AP collapse included complete collapse of the tongue base, as well as AP collapse of the velopharynx or epiglottis.

To account for multilevel collapse, patients with complete tongue base collapse were categorized as having AP collapse regardless of other collapse sites. Patients with lateral collapse in the absence of complete tongue base collapse were categorized as lateral, irrespective of velopharyngeal or epiglottic collapse pattern. These groupings were mutually exclusive. Patients demonstrating only partial collapse across sites were categorized separately as a partial collapse group (n = 178).

### Outcome Definitions

HGNS efficacy was assessed as the percent reduction in AHI from baseline to post-treatment evaluation.

Treatment success was defined using the Sher15 criterion^22^, consisting of a ≥50% reduction in AHI to <15 events per hour, consistent with prior HGNS outcome studies^13^.

### Statistical Analysis

Baseline characteristics and HGNS-only outcomes are reported as median values with interquartile ranges. HGNS-only outcomes excluded patients who underwent upper airway surgery (functional expansion pharyngoplasty, tonsillectomy, or partial uvulectomy) either prior to or concomitant with HGNS implantation without repeat DISE (n = 48). Outcomes were summarized within categories of AP-directed collapse, laterally directed collapse, and partial collapse, with further stratification by BMI using a median split (median BMI: 27.5 kg/m^2^).

The primary analysis tested the hypothesis that greater BMI is associated with a greater reduction in HGNS efficacy among patients with laterally directed collapse compared with those with AP-directed collapse. Multivariable regression was used to model HGNS efficacy as a continuous outcome, with BMI (continuous), lateral collapse pattern (binary), and their interaction as primary predictors. Models were adjusted for baseline AHI, a partial collapse × BMI interaction, surgical center, type of follow-up sleep study (tPSG vs home sleep test), and prior or concomitant upper airway surgery.

Secondary analyses repeated the primary model using absolute change in AHI as the outcome. Additional analyses evaluated treatment success as a dichotomous outcome using logistic regression. Sensitivity analyses using alternative definitions of treatment success (≥50% AHI reduction to <10 events per hour^23,24^ and ≥70% AHI reduction^25^) were also performed. All models were additionally adjusted for age and sex, with these results reported in the supplement. Statistical significance was defined as a two-sided P value < 0.05 for the primary interaction term.

## RESULTS

### Cohort Characteristics and Unadjusted Outcomes

Baseline characteristics for the pooled cohort, as well as stratified by cohort and pharyngeal collapse pattern, are summarized in Tables 1 and 2. HGNS efficacy outcomes overall, by cohort, and stratified by collapse pattern and body mass index (BMI) are summarized in Table 3. Baseline demographic and clinical characteristics were broadly similar between cohorts (Table 1). Patients with laterally directed collapse exhibited slightly higher baseline apnea–hypopnea index (AHI) and BMI compared with those with anteroposterior (AP) collapse patterns (Table 2).

**Table 1.**
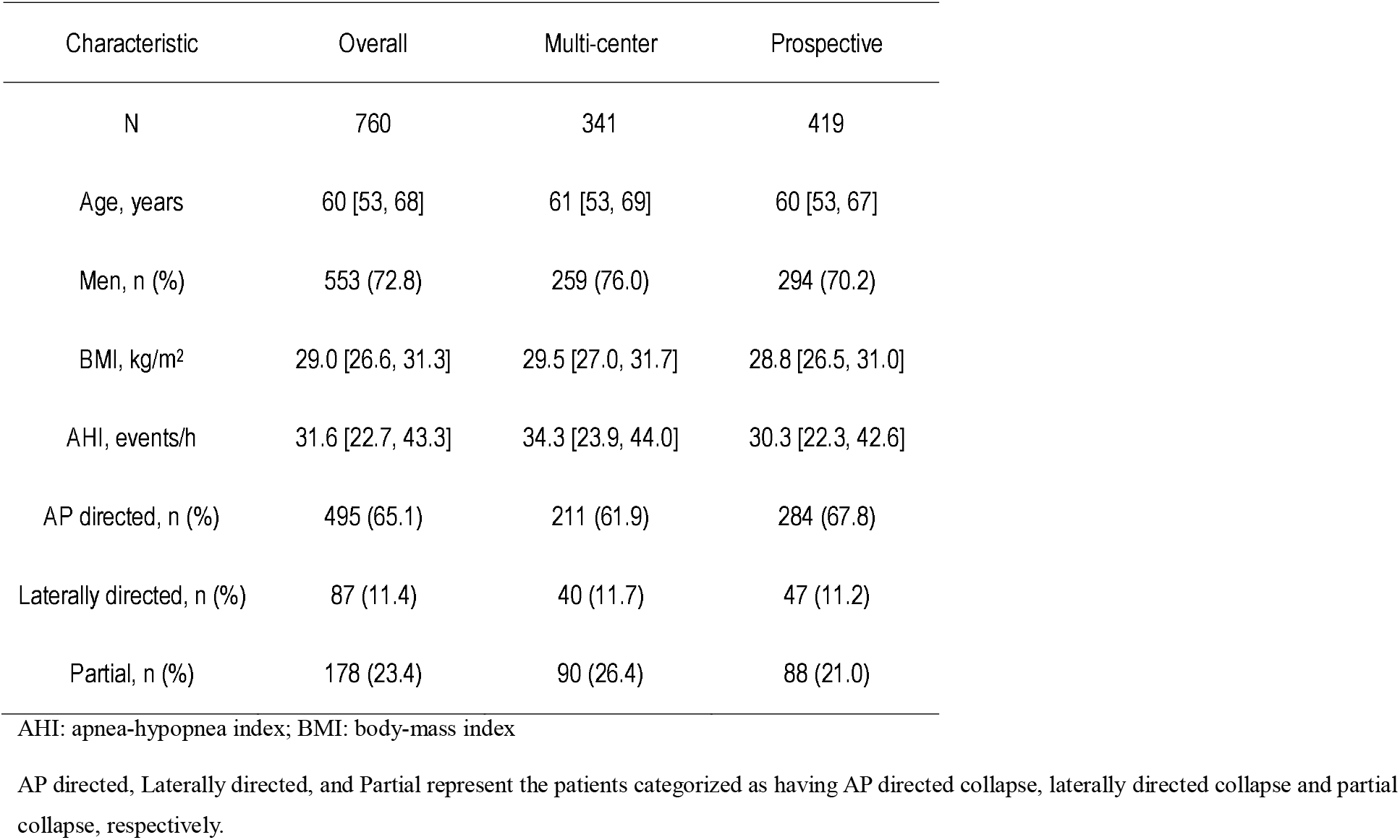
Baseline characteristics of the overall sample, across cohorts, and across pharyngeal collapse patterns.

| Characteristic | Overall | Multi-center | Prospective |
| --- | --- | --- | --- |
| N | 760 | 341 | 419 |
| Age, years | 60 [53, 68] | 61 [53, 69] | 60 [53, 67] |
| Men, n (%) | 553 (72.8) | 259 (76.0) | 294 (70.2) |
| BMI, kg/m<sup>2</sup> | 29.0 [26.6, 31.3] | 29.5 [27.0, 31.7] | 28.8 [26.5, 31.0] |
| AHI, events/h | 31.6 [22.7, 43.3] | 34.3 [23.9, 44.0] | 30.3 [22.3, 42.6] |
| AP directed, n (%) | 495 (65.1) | 211 (61.9) | 284 (67.8) |
| Laterally directed, n (%) | 87 (11.4) | 40 (11.7) | 47 (11.2) |
| Partial, n (%) | 178 (23.4) | 90 (26.4) | 88 (21.0) |

AHI: apnea-hypopnea index; BMI: body-mass index
AP directed, Laterally directed, and Partial represent the patients categorized as having AP directed collapse, laterally directed collapse and partial collapse, respectively.

**Table 2.**
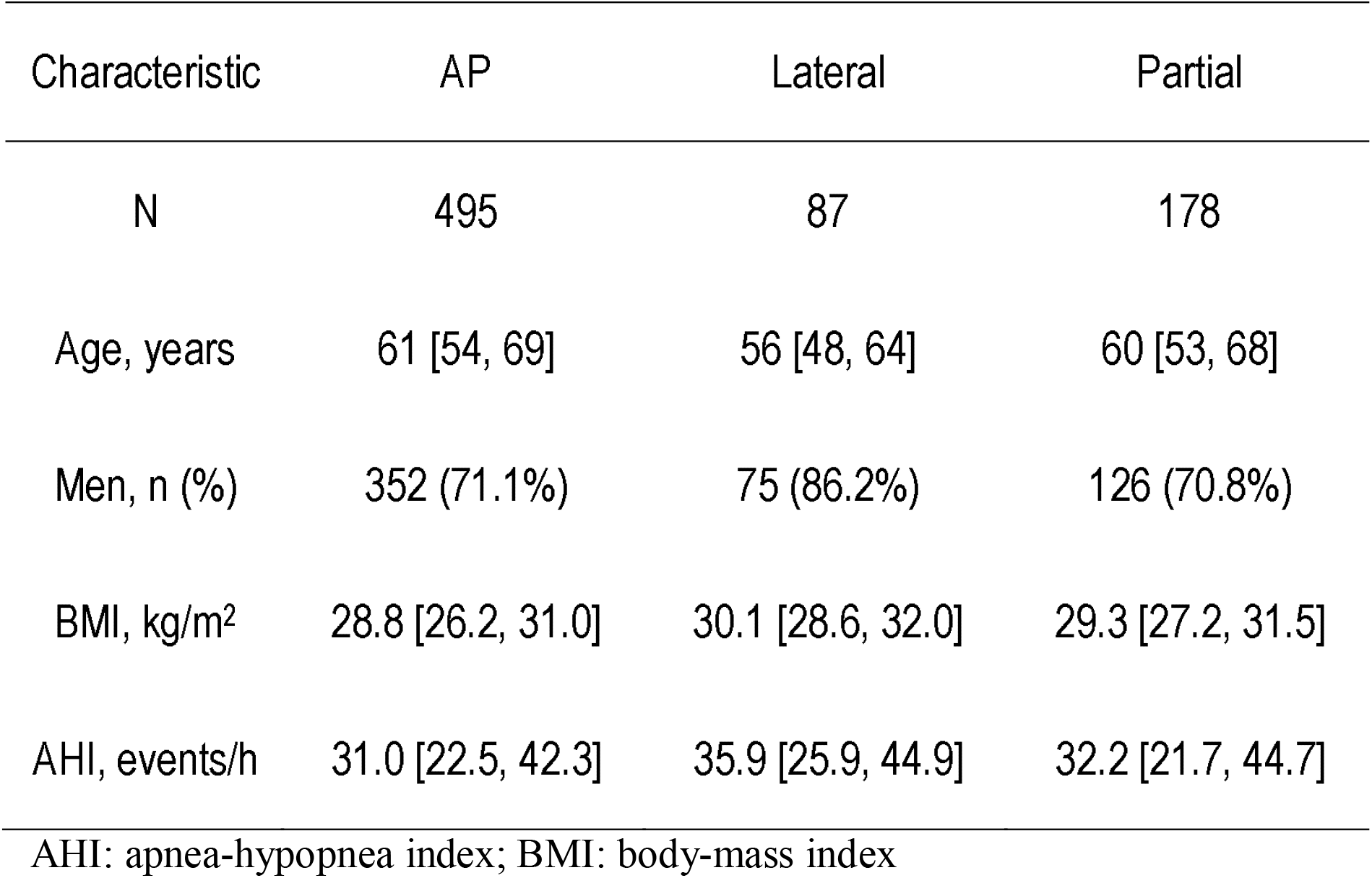
Baseline characteristics across anteroposterior (AP), lateral and partial collapse patterns.

**Table 3.** Hypoglossal nerve stimulation outcomes in pooled and individual cohorts, stratified by pharyngeal collapse pattern (anteroposterior, lateral, and partial).

| Cohort | Parameter | All | AP | Lateral | Partial |
| --- | --- | --- | --- | --- | --- |
| Pooled | N | 712 | 482 | 60 | 170 |
|  | Percent reduction AHI | 84.7 [62.6, 94.8] | 85.1 [64.1, 95.3] | 68.8 [43.8, 88.8] | 85.7 [65.9, 95.8] |
|  | N Success (%) | 547 (76.8%) | 377 (78.2%) | 34 (56.7%) | 136 (80.0%) |
| Multi-center | Percent reduction AHI | 80.9 [55.5, 92.6] | 81.6 [57.3, 93.4] | 67.9 [42.0, 89.4] | 80.8 [61.4, 93.9] |
|  | N Success (%) | 244 (71.6%) | 153 (72.5%) | 23 (57.5%) | 68 (75.6%) |
| Prospective | Percent reduction AHI | 87.4 [70.5, 96.0] | 87.7 [72.7, 96.3] | 70.0 [46.3, 87.0] | 89.1 [71.9, 96.5] |
|  | N Success (%) | 303 (81.7%) | 224 (82.7%) | 11 (55.0%) | 68 (85.0%) |

AHI: apnea-hypopnea index; BMI: body-mass index
AP directed, Laterally directed, and Partial represent the patients categorized as having AP directed collapse, laterally directed collapse and partial collapse, respectively.
Sample analyzed for this table only includes patients that underwent HGNS-only (n=712), i.e., excludes patients who underwent upper airway surgery (functional expansion pharyngoplasty, tonsillectomy, or partial uvulectomy) concomitant with or prior to HGNS without repeat DISE.

Across cohorts, HGNS efficacy and treatment success rates were lower among patients with laterally directed collapse compared with those with AP directed collapse (Table 3), consistent with prior reports^5,12–14^. Within the lateral and partial collapse groups, response rates were lower at higher BMI levels, whereas this pattern was less evident among patients with AP directed collapse.

### Primary Analysis: Interaction Between BMI and Collapse Pattern

In multivariable regression models evaluating HGNS efficacy as percent reduction in AHI, a statistically significant interaction was observed between BMI and the presence of lateral collapse (β [95% CI]: −15.9 [−30.0, −1.8]; Table 4). Stratified estimates indicated a steeper decline in HGNS efficacy with increasing BMI among patients with lateral collapse compared with those with AP collapse. Specifically, each 5 kg/m^2^ increase in BMI was associated with a −19.7 [−33.2, −6.2]% reduction in efficacy in patients with lateral collapse, compared with a −3.8 [−8.0, 0.36]% reduction in patients with AP collapse.

**Table 4.** Summary of the interaction between BMI and collapse pattern (lateral vs. AP) and the estimates (β coefficients or odds ratios) for the effect of BMI on HGNS outcomes within patients with lateral and AP collapse.

| Outcome | Interaction [95%CI], p-value | Lateral<br>Estimate [95%CI] | AP<br>Estimate [95%CI] |
| --- | --- | --- | --- |
| Percent reduction in AHI | -15.9 [-30.0, -1.8], p=0.027 | -19.7 [-33.2, -6.2] | -3.8 [-8.0, 0.36] |
| Absolute reduction in AHI | -5.2 [-10.2, -0.17], p=0.043 | -6.4 [-11.1, -1.6] | -1.2 [-2.7, 0.30] |
| Success | 4.1 [1.2, 14.0], p=0.023 | 1.1 [0.75, 1.5] | 4.4 [1.4, 14.3] |

AHI: apnea-hypopnea index; BMI: body-mass index.
Linear regression was used for continuous outcomes (percent and absolute AHI reduction) and logistic regression for binary outcomes (success). All models were adjusted for baseline AHI, the presence of partial collapse × BMI interaction, surgical center, type of follow-up sleep study, and previous or concomitant upper airway surgery.
Estimates for percent and absolute AHI reduction represents the additional reduction in HGNS efficacy outcome per 5 kg/m<sup>2</sup> increase in BMI
For the outcome 'Success,' estimates represent odds ratios for achieving treatment success ( $\geq 50\%$ AHI reduction to $<10$ events/h) per 5 kg/m<sup>2</sup> increase in BMI.
The interaction represents the additional reduction in HGNS efficacy outcomes per 5 kg/m<sup>2</sup> increase in BMI that is observed in patients with lateral collapse compared to those with AP collapse.

When absolute change in AHI was used as the outcome, a similar BMI × collapse pattern interaction was observed (−5.2 [−10.2, −0.17] events/h; Table 4). Increasing BMI was associated with a greater loss of efficacy in patients with lateral collapse (−6.4 [−11.1, −1.6] events/h per 5 kg/m^2^ increase in BMI) compared with those with AP collapse (−1.2 [−2.7, 0.30] events/h per 5 kg/m^2^ increase in BMI).

### Secondary Analyses: Treatment Success

In logistic regression models evaluating treatment success, defined as a ≥50% reduction in AHI to <15 events per hour, the interaction between BMI and laterally directed collapse remained statistically significant (odds ratio [95% CI]: 4.1 [1.2, 14.0]; Table 4). Increasing BMI was associated with reduced odds of treatment success among patients with lateral collapse, whereas BMI was not significantly associated with treatment success among patients with AP collapse. Results were consistent when alternative definitions of treatment success were applied (Table E1, Online Supplement).

### Partial Collapse Analyses

Because the primary model included an interaction term between partial collapse and BMI, the association between BMI and HGNS efficacy in patients with partial collapse could also be evaluated. In this group, HGNS efficacy decreased moderately with increasing BMI (−9.1% [−15.4, −3.3%] per 5 kg/m^2^ increase in BMI).

In supplemental analyses, patients with partial collapse demonstrated higher HGNS efficacy compared with those with lateral collapse, with an improvement of 8.9% [1.8, 15.2%] in percent AHI reduction. This difference was associated with a higher likelihood of treatment success (odds ratio [95% CI]: 3.3 [1.6, 6.5]). These findings were comparable to the differences observed between AP and lateral collapse patterns (8.5% [2.0, 14.4%] improvement and 2.7-fold [1.5, 5.1] higher likelihood of success, respectively).

### Additional Analyses

All primary findings were similar after additional adjustment for age and sex (Table E2, Online Supplement). Age and sex did not demonstrate significant interactions with collapse pattern. When collapse sites were examined individually, the association between BMI and HGNS efficacy within each site mirrored that of the broader collapse category to which they were assigned (Table E3, Online Supplement). Specifically, increasing BMI had minimal impact on HGNS efficacy in patients with complete AP velum, tongue base, or epiglottic collapse, whereas efficacy declined with increasing BMI in patients with complete OLW collapse and CCC.

## DISCUSSION

In this pooled analysis of two independent HGNS cohorts, we observed that the relationship between body mass index (BMI) and HGNS efficacy differed according to pharyngeal collapse pattern. Specifically, increasing BMI was associated with progressively reduced HGNS efficacy in patients with lateral collapse patterns, whereas BMI had comparatively little impact on treatment efficacy among patients with predominantly anteroposterior (AP) collapse. These findings indicate that the effect of BMI on HGNS outcomes is not uniform across collapse patterns and suggest that BMI and collapse site should be considered jointly when evaluating treatment response.

Previous studies have demonstrated that HGNS efficacy and treatment success decline with increasing BMI^10,26–28^. The present study extends these observations by demonstrating that this association is modified by the site and pattern of pharyngeal collapse. Among patients with lateral collapse, a 5 kg/m^2^ increase in BMI was associated with a marked reduction in HGNS efficacy and a substantial decrease in the likelihood of treatment success. In contrast, among patients with AP collapse, increasing BMI was associated with only modest changes in efficacy and was not associated with reduced odds of treatment success. Together, these findings suggest that BMI functions as a risk factor for reduced HGNS response primarily in the presence of lateral collapse.

Importantly, the observed interaction also indicates that lateral collapse does not uniformly preclude favorable HGNS outcomes^5,12,14^. In patients with lower BMI, HGNS efficacy and success rates were similar between AP and lateral collapse patterns. This observation suggests that lateral collapse patterns commonly considered difficult to treat may remain amenable to HGNS in the setting of lower BMI.

Several mechanisms may explain why increasing obesity preferentially impairs HGNS efficacy in patients with lateral walls collapse compared to those with AP collapse. First, increased obesity is associated with greater volume of the parapharyngeal fat pads and adipose tissue in the base of the tongue^29–31^, and is a well-established contributor to increased pharyngeal collapsibility^15,17^. Accordingly, we suggest that when the laterally collapsing airway is only mildly collapsible, as is more common in patients with lower BMI, AP directed tongue movement can generate sufficient traction to promote lateral airway opening. This mechanical coupling between the tongue and pharyngeal tissues (i.e., glossopharyngeal coupling) may play a role in mediating the effects of HGNS on lateral collapse; however, with greater obesity and collapsibility, this coupling may be compromised. In contrast, for patients with AP collapse, increased adiposity may still increase collapsibility, but the airway remains treatable with HGNS, as the therapy directly targets the tongue base.

In addition, obesity—particularly central adiposity—is associated with reduced lung volumes and diminished caudal traction on the upper airway^32–35^. Reduced lung volume–related traction may further impair the ability of HGNS-induced tongue movement to influence lateral airway dimensions, thereby amplifying the adverse effect of increasing BMI in patients with lateral collapse. While these mechanisms are consistent with established physiological principles, further studies incorporating direct pattern of upper airway collapse, measures of upper airway collapsibility, and lung volume are needed to clarify their relative contributions.

Although not a primary focus of the study, analyses of patients with partial collapse provide additional context. In this group, HGNS efficacy declined moderately with increasing BMI, but overall treatment response remained favorable and comparable to that observed in patients with complete AP collapse. Partial collapse was associated with higher efficacy and success rates than lateral collapse, and these differences were similar in magnitude to those observed between AP and lateral collapse. Furthermore, the directional pattern of partial collapse (AP versus lateral) did not significantly influence HGNS efficacy. These findings suggest that partial collapse represents a collapse phenotype that is generally responsive to HGNS, with modest susceptibility to BMI-related reductions in efficacy.

Several limitations should be considered when interpreting these findings. First, the protocol for scoring DISE data differed between the retrospective and prospective cohorts. In the retrospective cohort, collapse patterns were assigned based on consensus scoring by multiple blinded reviewers^5^, whereas in the prospective cohort, scoring was performed clinically at the time of DISE. However, effect sizes for the BMI × collapse interaction were similar across cohorts, and test–retest analyses in the retrospective cohort demonstrated good agreement for identification of complete lateral and complete AP collapse, suggesting that differences in DISE scoring approach did not materially affect the results (Online Supplement).

Second, treatment efficacy was primarily assessed using titration polysomnography, which estimates AHI during periods of optimal stimulation and may overestimate real-world efficacy compared with single-setting home sleep apnea testing^3^. However, titration studies are performed under controlled conditions, minimize confounding due to changes in body weight or therapy adherence, and reduce discontinuation bias. Although absolute efficacy estimates may be higher than those obtained from home testing, we do not expect this approach to influence the observed interaction between BMI and collapse pattern. Consistent with this, sensitivity analyses adjusting efficacy estimates to approximate home sleep testing values resulted in a uniform downward shift without altering the BMI-related differences across collapse patterns (Online Supplement).

Third, the number of patients with BMI values above 35 kg/m^2^ was limited, restricting inference regarding HGNS efficacy at higher BMI levels. Larger studies will be needed to evaluate whether the observed interaction persists across the full range of BMI currently considered eligible for HGNS therapy.

Finally, sleep study scoring criteria were not uniform across centers in the multicenter cohort, which may introduce variability in absolute AHI values. However, because scoring definitions were consistent within individuals across baseline and follow-up studies, percent AHI reduction is unlikely to be substantially affected, and the interaction between BMI and collapse pattern were consistent between the cohorts (Online Supplement).

In summary, this study demonstrates that BMI modifies HGNS efficacy in a collapse pattern–specific manner, with increasing BMI associated with reduced treatment response in patients with lateral pharyngeal collapse but not in those with AP collapse. These findings underscore the importance of considering anatomical collapse patterns alongside BMI when evaluating HGNS outcomes and provide a framework for more refined assessment of factors influencing treatment response.

## Data Availability

The data supporting the findings of this study are not publicly available due to privacy and regulatory restrictions, but are available from the corresponding author upon reasonable request and with appropriate institutional approvals.

## Author Contributions

Conception and design of the work: DV, SAS, PH

Data collection/management: DV, JLGCM, EK, PH, DK, MD

Statistical analysis: DV, SAS, TC

Data interpretation: DV, EK, AW, DK, MD, JLGCM, AA, SAS, PH

Drafting the article: DV, SAS, PH

Critical revision of the article: DV, EK, AW, DK, MD, JLGCM, TC, AA, SAS, PH

Final approval of the version to be published: DV, EK, AW, DK, MD, JLGCM, AA, TC, DPW, SAS, PH

