## Supplementary material for "Differential association of body mass index with hypoglossal nerve stimulation efficacy by pharyngeal collapse pattern in obstructive sleep apnea": Online Supplement

**Association between body mass index and unilateral hypoglossal nerve stimulation efficacy depends on the pattern of pharyngeal collapse in obstructive sleep apnea**

Daniel Vena^1^, Eric J. Kezirian^2^, Andrew Wellman^1^, David Kent^3^, Mark D’Agostino^4,5^, Joao L.G.C. Monteiro^1^, Azarbarzin A^1^, Tom Chen^6^, Ludovico Messineo^1^, Neda Esmaeili^1^, Scott A. Sands^1^*, Phillip Huyett^7^*.

^1^Division of Sleep and Circadian Disorders, Brigham and Women's Hospital and Harvard Medical School, Boston, MA.

^2^Department of Head and Neck Surgery, David Geffen School of Medicine at UCLA, Los Angeles, California.

^3^Department of Otolaryngology-Head and Neck Surgery, Vanderbilt University Medical Center, Nashville, Tennessee.

^4^Southern New England Ear, Nose and Throat Group, Middlesex Hospital, Middlesex, Connecticut.

^5^Department of Surgery, Yale University School of Medicine, New Haven, Connecticut.

^6^Department of Population Medicine, Harvard Medical School and Harvard Pilgrim Health Care Institute, Boston, MA

^7^Department of Otolaryngology, Massachusetts Eye and Ear, Boston, MA

*_,_^†^Equal contributions

**Correspondence:**

Daniel Vena

221 Longwood Ave., Boston MA, 02115

Division of Sleep and Circadian Disorders, Department of Medicine, Brigham & Women's Hospital & Harvard Medical School, Boston, Massachusetts.

**Interaction between BMI and pharyngeal collapse pattern on HGNS success using alternative definitions of success**

To demonstrate consistency of the interaction effect between BMI and lateral collapse on HGNS success, we reran our secondary analysis with other commonly used definitions of success: 50% AHI reduction to <10 /h^1,2^ (category 2), 70% AHI reduction^3^ (category 3), and 50% AHI reduction to <5 /h. Results, summarized in Table E1 were consistent with the HGNS success criterion used in the main paper (Category 1: 50% AHI reduction to <15 /h). The interaction was more modest when success was defined as post-treatment AHI <5 events/h, although the overall pattern remained directionally consistent with the other success definitions.

Table E1. Interaction between BMI and laterally directed collapse on HGNS success defined across commonly used definitions of success

| HGNS success category | Interaction | AP-directed | Laterally-directed |
| --- | --- | --- | --- |
| Category 1 (main paper) | 4.1 [ 1.2, 14.0 ] | 1.1 [ 0.75, 1.5 ] | 4.4 [ 1.4, 14.3 ] |
| Category 2 | 2.6 [ 0.86, 7.9 ] | 1.2 [ 0.79, 1.9 ] | 3.1 [ 1.1, 9.1 ] |
| Category 3 | 3.3 [ 1.0, 10.4 ] | 1.4 [ 0.99, 1.9 ] | 4.5 [ 1.5, 13.7 ] |
| Category 4 | 1.4 [0.46, 4.0] | 1.6 [1.2, 2.2] | 2.2 [0.78, 6.3] |

**Adjustment of age and sex**

The analysis evaluating the interaction between lateral collapse and BMI on HGNS efficacy, as described in the main manuscript, was repeated with adjustment for age and sex. Adjustment had no meaningful impact on the interaction term or on the estimates of BMI effects within the lateral and AP collapse groups (Table E2).

Table E2. Summary of the interaction between BMI and collapse pattern (lateral vs. AP) and adjusted estimates (β coefficients or odds ratios) for the effect of BMI on HGNS outcomes within lateral and AP collapse groups, with models adjusted for age and sex.

| Outcome | Interaction [95%CI], p-value | Lateral Estimate [95%CI] | AP Estimate [95%CI] |
| --- | --- | --- | --- |
| Percent reduction in AHI | −15.6 [−29.7, −1.6], p=0.029 | −19.4 [−32.8, −5.9] | −3.7 [−7.8, 0.42] |
| Absolute reduction in AHI | −5.1 [−10.1, −0.1], p=0.043 | −6.3 [−11.1, −1.5] | −1.2 [−2.6, 0.32] |
| Success | 4.1 [1.2, 14.1], p=0.023 | 4.3 [1.3, 14.0] | 1.0 [0.73, 1.5] |

AHI: apnea-hypopnea index; BMI: body-mass index.

Linear regression was used for continuous outcomes (percent and absolute AHI reduction) and logistic regression for binary outcomes (success). All models were adjusted for age, sex, baseline AHI, the presence of partial collapse × BMI interaction, surgical center, type of follow-up sleep study, and previous or concomitant upper airway surgery.

Estimates for percent and absolution AHI reduction represents the additional reduction in HGNS efficacy outcome per 5 kg/m^2^ increase in BMI

For the outcome 'Success,' estimates represent odds ratios for achieving treatment success (≥50% AHI reduction to <15 events/h) per 5 kg/m² increase in BMI..

The interaction represents the additional reduction in HGNS efficacy outcomes per 5 kg/m² increase in BMI that is observed in patients with lateral collapse compared to those with AP collapse.

**Age and sex interaction with BMI on HGNS efficacy**

To examine whether age and sex were differentially associated with HGNS efficacy in patients with AP versus lateral collapse, we repeated the primary analysis from the main paper, substituting age and sex for BMI. Specifically, we conducted a multivariable mixed model regression to evaluate HGNS efficacy as a function of the presence of lateral collapse, age/sex and their interaction (age/sex × collapse pattern), and adjusted for covariates baseline AHI, the presence of partial collapse, surgical center, type of follow-up sleep study, and previous or concomitant upper airway surgery.

The interaction between age and laterally directed collapse was not statistically significant in the linear regression model evaluating HGNS efficacy (β [95% CI]: 6.2 [−0.64, 7.9]% per 10-year increase in age; p = 0.08). Nonetheless, there was a suggestive trend toward modestly greater efficacy with increasing age among patients with lateral collapse (5.1 [−1.2, 11.5]% per 10-year increase), a pattern not observed in patients with AP collapse (−1.1 [−3.5, 1.4]% per 10-year increase). Similarly, the interaction between sex and laterally directed collapse was not statistically significant (β [95% CI]: −3.1 [−23.9, 17.6]%; p = 0.76 for men compared with women).

**Interaction between BMI and various pharyngeal site-of-collapse categories in predicting HGNS efficacy**

We also evaluated the interaction between various combinations of different site-of-collapse categories (e.g., oropharyngeal lateral wall collapse combined with anteroposterior velum collapse) and BMI in explaining HGNS efficacy. To achieve this, we divided the analysis into five logical components. In the first component (analysis group A in Table E3), we examined the individual categories that constitute laterally-directed collapse: complete concentric collapse of the soft palate (CCC) and complete oropharyngeal lateral wall collapse (OLW). Our goal was to determine whether patients within each category exhibited substantial reductions in HGNS efficacy, similar to the parent category of lateral collapse. Indeed, lateral collapse defined by CCC alone, OLW alone, or a combination of CCC and OLW demonstrated substantial reductions in HGNS efficacy with increasing BMI (Table E3), with effect sizes comparable to or greater than those reported in the main paper.

Since the findings for the different site-of-collapse categories within lateral collapse were similar to those for lateral collapse overall, we combined them back into a single category of lateral collapse. We then assessed whether anteroposterior (AP) velum and epiglottic collapse, when occurring concurrently with laterally-directed collapse, influenced its interaction with BMI in explaining HGNS efficacy; as well as how this compared to isolated lateral collapse (i.e., without any accompanying collapse patterns). As shown in analysis group B results from Table E3, isolated lateral collapse and lateral collapse combined with AP velum demonstrated substantial reductions in HGNS efficacy with increasing BMI, with effect sizes comparable to or greater than those reported in the main paper. We note that the combination of lateral collapse with epiglottic collapse could not be evaluated due to an insufficient number of patients exhibiting this pattern.

Altogether, the findings from Table E3 demonstrate that patients with lateral collapse—whether defined as CCC, OLW collapse, or a combination of the two, and whether occurring in isolation or concurrently with AP velum or epiglottic collapse—exhibit substantial reduction in HGNS efficacy as BMI increases.

Next, we performed a similar analysis for AP collapse patterns in analysis groups C, D, and E (Table E3) to demonstrate that HGNS efficacy remains relatively stable with increasing BMI across the individual site-of-collapse patterns that fall within the category of AP collapse. In analysis group C we evaluate change in HGNS efficacy with increasing BMI within various categories of tongue collapse: isolated tongue collapse, tongue collapse concurrent with collapse of the AP velum, epiglottis, and then AP-directed collapse without any tongue involvement for comparison. As summarized in Table E3, HGNS efficacy varies minimally (or even increases) with increasing BMI across the various categories, with effect sizes comparable to that of the main paper. We note that results from concurrent tongue and epiglottic collapse were not reliable given the small sample size.

In analysis group D, we evaluated the change in HGNS efficacy with increasing BMI across various categories of AP velum collapse (Table E3). Only isolated AP velum collapse had a sufficient sample size for meaningful conclusions, and it exhibited results consistent with AP-directed collapse, showing minimal change in HGNS efficacy with increasing BMI.

Lastly, analysis group E examined the change in HGNS efficacy with increasing BMI in cases of isolated epiglottic collapse. Although the sample size was small, the effect size fell between those observed for AP-directed and laterally-directed collapse.

Table E3. The effect of BMI on HGNS efficacy within various site-of-collapse categories.

| Analysis group | Collapse category | N | Difference in Efficacy per 5 unit increase in BMI.  Beta [95%CI] | VOTE scores comprising collapse category |
| --- | --- | --- | --- | --- |
| A | CCC without OLW | 10 | −63.8 [−132.1, −3.4] | V2c, V2cE2 |
|  | OLW without CCC | 72 | −15.3 [−30.4, −0.18] | O2, V2apO2, O2E2 |
|  | CCC + OLW | 5 | −17.6 [−62.1, 26.9] | V2cO2, V2cO2E2 |
| B | Isolated lateral collapse | 71 | −17.1 [−33.6, −0.65] | V2c, O2, V2cO2 |
|  | Lateral + AP velum | 15 | −17.2 [−44.2, 9.9] | V2apO2 |
|  | Lateral + Epiglottis | 1 | N/A | O2E2 |
| Analysis group | Collapse category | N | Change in Efficacy with BMI Beta [95%CI] | VOTE scores within collapse category |
| C | Isolated Tongue | 154 | −4.3 [−11.5, 2.9] | T2 |
|  | Tongue + AP velum | 201 | −4.4 [−10.8, 1.9] | V2apT2, V2apT2E2 |
|  | Tongue + Epiglottis | 2 | 60.3 [204.8, 69.5] | T2E2 |
|  | AP collapse without Tongue | 138 | 0.98 [−8.1, 10.1] | V2ap, V2apE2 |
| D | Isolated AP velum | 111 | 1.6 [−8.3, 11.6] | V2ap |
|  | AP velum + Epiglottis | 10 | −12.1 [−57.5, 33.3] | V2apE2 |
|  | AP collapse without AP velum | 10 | −5.2 [−10.0, −0.48] | T2, T2E2 |
| E | Isolated Epiglottis | 17 | −7.4 [−36.4, 21.7] | E2 |

Efficacy defined by the percent reduction in apnea-hypopnea index from baseline.

CCC: Complete concentric collapse of the soft palate; OLW: oropharyngeal lateral walls; AP: anteroposterior

**Effect of partial pharyngeal collapse on HGNS efficacy**

To study the effect of partial collapse on HGNS efficacy we used multivariable mixed model regression to evaluate HGNS efficacy (continuous dependent variable) as a function of the presence of partial pharyngeal collapse pattern (dichotomous independent variable) and AP-directed collapse (dichotomous), and adjusted for covariates baseline AHI (continuous), surgical center, type of follow-up sleep study (titration PSG or single-setting home sleep test), and previous or concomitant upper airway surgery (functional expansion pharyngoplasty, tonsillectomy, and partial uvulectomy). We repeated this analysis using successful treatment response as the outcome (dichotomous) and logistic regression instead of linear regression. These analyses quantified increased HGNS efficacy and success rate in patients with partial collapse relative to lateral collapse. For comparison, the increased HGNS efficacy and success in AP collapse was also analyzed and presented. As described in the main paper, HGNS efficacy in partial collapse was greater by 9.0 [0.33, 17.6]% over lateral collapse, with a corresponding 3-fold greater likelihood of a successful response (odds ratio [95% CI]: 3.3 [1.6, 6.5]), both comparable to the 8.5 [2.0, 14.4]% efficacy gain and 2.7-fold [1.5, 5.1] increased likelihood seen in AP collapse over lateral collapse.

We also evaluated the effect of increasing BMI on HGNS efficacy and success rate in patients with partial collapse. To do so, we used the same interaction model described in the main paper. The analysis showed that BMI had a moderate effect on HGNS outcomes, decreasing efficacy by −9.2 [−16.1, −2.3]% per 5 kg/m^2^. Success rate also decreased by 1.6 times per 5 kg/m^2^, but this did not reach statistical significance (odds ratio: 1.6 [0.83, 3.0]).

Lastly, we investigated the effect of pattern of *partial* collapse (i.e., AP vs. lateral) using a multivariable mixed model regression that evaluated HGNS efficacy as function of lateral *partial* collapse, adjusting out complete collapse, and adjusting for the same covariates described above. The coefficient for laterally-directed partial collapse quantifies the difference in HGNS efficacy of laterally-directed partial collapse compared to AP-directed partial collapse. The model showed that there was no significant difference in HGNS efficacy (4.8 [−6.5, 16.0]%) and success rate (odds ratio [95%CI]: 1.5 [0.55, 4.2]) between lateral and AP partial collapse. Overall, this suggests that the pattern of collapse is less important in the context of partial collapse.

**Interaction between BMI and laterally directed collapse on HGNS outcomes, analyzed separately by cohort**

We report the interaction between BMI and the presence of lateral collapse on HGNS efficacy and success rate separately for the multi-center and prospective cohorts. The interaction effect for HGNS efficacy was substantial in both cohorts, measuring −13.7% (95% CI: −31.6, 4.1) in the prospective cohort and somewhat larger in the multi-center cohort at −18.7% (95% CI: −41.1, 3.6). Similarly, the interaction effect for HGNS success rate (odds ratio [95%CI]) in the multi-center cohort was 3.4 [0.77, 14.7], and a comparable effect was found in the prospective cohort at 3.9 [0.33, 28.9]. These findings suggest that single-reviewer scoring yielded consistent relationships between site of collapse, BMI, and HGNS efficacy, supporting its reliability for this analysis.

**Inter-rater reliability for classifying pharyngeal collapse as lateral or anteroposterior**

We assessed inter-rater reliability for categorizing airway collapse as lateral or not and as AP or not. To do so, we fit a logistic mixed model with random effects for video and rater (model equation: $\mathrm{logit}P\left( Y_{ij}=1 | \eta_{i},\zeta_{j} \right)=\alpha+\eta_{i}+\zeta_{j}$ for the $i$th video and $j$th rater). The logistic-scale within-task intraclass correlation coefficient (ICC) was calculated based on the variance of the random effects $\sigma_{\eta}^{2},\sigma_{\zeta}^{2}$using the equation^4^: $\mathrm{ICC}_{\mathrm{Task}}=\frac{\sigma_{\eta}^{2}}{\sigma_{\eta}^{2}+\sigma_{\zeta}^{2}+\pi^{2}/3}$. The model demonstrated a high ICC for lateral collapse (0.72) and a fairly strong ICC for AP collapse (0.50), indicating good agreement among raters for both categorizations. These findings again support the reliability of site-of-collapse assessments made by a single reviewer in this analysis.

**Interaction between BMI and lateral collapse on HGNS efficacy adjusted to reflect efficacy from home sleep apnea test.**

To address the limitation that HGNS efficacy was determined based on optimal setting AHI, which overestimates actual HGNS efficacy^5^, we estimated HGNS efficacy as it would be if treatment AHI were measured using an HSAT. We leveraged data from patients who underwent a post-treatment HSAT (n=60) to compute the mean and standard deviation of HSAT-derived HGNS efficacy distribution. We subsequently applied a linear transformation to adjust the distribution of HGNS efficacy calculated based on the titration study at optimal stimulation to match the mean and standard deviation of HGNS efficacy derived from the HSAT. With the adjusted HGNS efficacy, we re-ran our primary multivariable regression analysis, evaluating estimated HSAT-derived HGNS efficacy as a function of laterally-directed pharyngeal collapse, BMI, and their interaction (BMI × collapse pattern), adjusting for the same covariates as in the main analysis.

This adjustment resulted in a uniform ~10% downward shift in overall efficacy (Figure E1). The model again identified a significant interaction between BMI and lateral collapse in predicting HGNS efficacy (β [95% CI]: −15.6 [−29.3, −1.9]%, p = 0.026). Specifically, lateral collapse was associated with a similar reduction in efficacy (−19.2 [−32.4, −6.1]% per 5 kg/m² increase in BMI) compared to AP collapse (−3.6 [−7.7, 0.41]% per 5 kg/m² increase in BMI) (Figure E1).

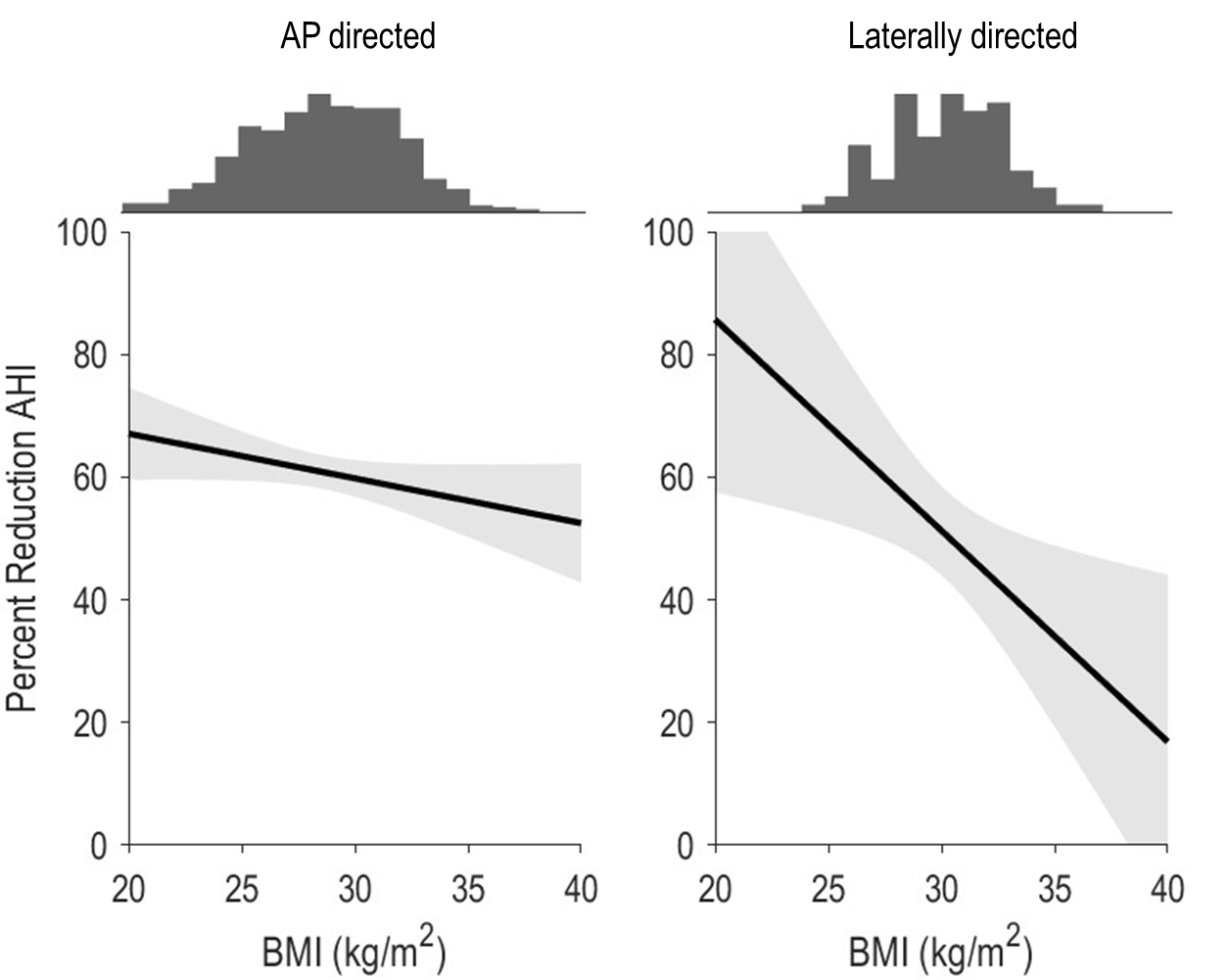

Figure E1. Impact of body mass index (BMI) on adjusted hypoglossal nerve stimulation (HGNS) efficacy (percent reduction AHI) across AP- and laterally directed collapse patterns. Histograms at the top of the figure illustrates the BMI distribution within each pattern of collapse. HGNS efficacy was adjusted to reflect efficacy determined from follow-up home sleep testing.

**Differences in prevalences of collapse patterns across individual surgical centers**

We assessed variability in collapse prevalence across individual centers from the multi-center cohort using chi-square testing and quantified the magnitude of between-center differences using Cramér’s V, where values <0.10 indicate trivial differences, 0.10–0.30 indicate small differences, 0.30–0.50 indicate moderate differences, and >0.50 indicate large differences. Across the nine individual centers of the multi-center cohort, most collapse categories demonstrated only small between-center differences (Table 3 and Table 4), including lateral collapse (V=0.19), partial collapse (V=0.28), velum AP collapse (V=0.23), oropharyngeal lateral wall collapse (V=0.21), and epiglottic collapse (V=0.17). Moderate between-center differences were observed only for AP collapse (V=0.34) and tongue-base collapse (V=0.32). Because all available DISE videos were centrally re-scored using standardized VOTE criteria and only consensus classifications were retained, these differences are unlikely to reflect variability in collapse scoring itself. Although some variability could reflect differences in DISE technique across centers (e.g., sedative agent or sedation depth), we suspect that the larger contributor was center-level variation in patient selection and HGNS candidacy practices.

Table 3. Prevalence of different collapse patterns across individual sites in the multicenter cohort.

| Center | N | Lateral | AP | Partial | V2ap | O2 | T2 | E2 |
| --- | --- | --- | --- | --- | --- | --- | --- | --- |
| 1 | 43 | 20.93 | 41.86 | 37.21 | 23.26 | 16.28 | 18.60 | 4.65 |
| 2 | 14 | 7.14 | 64.29 | 28.57 | 64.29 | 7.14 | 35.71 | 0.00 |
| 3 | 35 | 17.14 | 40.00 | 42.86 | 20.00 | 2.86 | 25.71 | 0.00 |
| 4 | 42 | 7.14 | 61.90 | 30.95 | 42.86 | 7.14 | 50.00 | 4.76 |
| 5 | 33 | 12.12 | 57.58 | 30.30 | 27.27 | 15.15 | 45.45 | 3.03 |
| 6 | 89 | 5.62 | 85.39 | 8.99 | 46.07 | 4.49 | 60.67 | 10.11 |
| 7 | 18 | 22.22 | 50.00 | 27.78 | 33.33 | 22.22 | 33.33 | 0.00 |
| 8 | 27 | 18.52 | 62.96 | 18.52 | 40.74 | 18.52 | 48.15 | 3.70 |
| 9 | 28 | 7.14 | 50.00 | 42.86 | 42.86 | 7.14 | 21.43 | 3.57 |

Table 4. Statistical estimates describing differences across sites.

| Site of collapse | P-value | Cramers V | Interpretation |
| --- | --- | --- | --- |
| Lat2cat | 0.133 | 0.194 | Small |
| AP2cat | <0.0001 | 0.340 | Moderate |
| Partial | 0.001 | 0.285 | Small |
| Vap2cat | 0.021 | 0.234 | Small |
| O2cat | 0.080 | 0.207 | Small |
| T2cat | <0.0001 | 0.320 | Moderate |
| E2cat | 0.330 | 0.167 | Small |

Encouragingly, when individual centers of the multi-center cohort were pooled, these center-level differences appeared to average out, resulting in collapse distributions that were generally similar to the prospective cohort (Table 5). Collapse distributions were broadly similar between cohorts, particularly for the mutually exclusive categories studied in the current paper (lateral, AP, and partial collapse). Lateral collapse prevalence was nearly identical between cohorts (11.7% vs. 11.2%; Cramér’s V=0.008), while differences in AP collapse (61.9% vs. 67.8%; V=0.062) and partial collapse (26.4% vs. 21.0%; V=0.063) were trivial. The main difference between cohorts was a higher prevalence of complete AP velum collapse in the prospective cohort (50.4% vs. 37.0%; V=0.134), representing a small difference. Differences in tongue-base collapse (50.4% vs. 42.8%; V=0.075) and oropharyngeal lateral wall collapse (14.3% vs. 9.7%; V=0.070) were trivial. Overall, these findings support the consistency of the primary collapse classifications between cohorts despite differences in DISE acquisition. We note that complete concentric velum collapse was not compared between cohorts because this pattern was only systematically re-scored in the multi-center cohort.

Consistent with this, the primary interaction between BMI and lateral collapse was similar in magnitude across the multi-center and prospective cohorts, suggesting that the relationship between obesity and collapse pattern was robust despite center-level differences. Specifically, the interaction effect for HGNS efficacy was substantial in both cohorts, measuring −13.7% (95% CI: −31.6, 4.1) in the prospective cohort and somewhat larger in the multicenter cohort at −18.7% (95% CI: −41.1, 3.6). Similarly, the interaction effect for HGNS success rate was comparable between cohorts, with odds ratios (95% CI) of 3.4 (0.77, 14.7) in the multicenter cohort and 3.9 (0.33, 28.9) in the prospective cohort. Together, these findings suggest that the relationships between BMI, collapse pattern, and HGNS outcomes were consistent across cohorts despite differences in prevalence of collapse patterns and DISE acquisition protocols.

Table 5. Prevalence of collapse patterns between multicenter and prospective cohorts.

| Site of Collapse | Multicenter Cohort | Prospective Cohort | P-value | Absolute Difference (%) | Cramers V |
| --- | --- | --- | --- | --- | --- |
| Lateral | 40/341 (11.7%) | 47/419 (11.2%) | 0.825 | -0.51 | 0.008 |
| AP | 211/341 (61.9%) | 284/419 (67.8%) | 0.089 | 5.90 | 0.062 |
| Partial | 90/341 (26.4%) | 88/419 (21.0%) | 0.081 | -5.39 | 0.063 |
| V2ap | 126/341 (37.0%) | 211/419 (50.4%) | 0.000 | 13.41 | 0.134 |
| O2 | 33/341 (9.7%) | 60/419 (14.3%) | 0.052 | 4.64 | 0.070 |
| T2 | 146/341 (42.8%) | 211/419 (50.4%) | 0.038 | 7.54 | 0.075 |
| E2 | 16/341 (4.7%) | 20/419 (4.8%) | 0.958 | 0.08 | 0.002 |

**BMI threshold sensitivity analysis of HGNS efficacy by collapse pattern**

Differences in HGNS efficacy between high and low BMI groups were consistent across BMI thresholds of 30, 32, and 35 kg/m² (Table 6,Table 7, and Table 8). In patients with AP collapse, HGNS efficacy was relatively stable regardless of whether BMI cutoffs of 30, 32, or 35 kg/m² were used. In contrast, among patients with lateral collapse, higher BMI categories were consistently associated with lower treatment response rates and lower percent reduction in AHI. Results using a BMI cutoff of 30 kg/m² were very similar to the median split presented in the manuscript (Table 6). With a cutoff of 32 kg/m², the reduction in efficacy appeared shifted toward lower BMI values, such that even the “low BMI” lateral collapse group had somewhat reduced efficacy compared with AP collapse (Table 7). At the BMI >35 kg/m² threshold, there were very few patients with lateral collapse (n=2), but both had poor outcomes (Table 8). However, because only 12 patients overall and only 2 patients with lateral collapse had BMI >35 kg/m², these findings should be interpreted cautiously.

Table 6. Hypoglossal nerve stimulation outcomes in the pooled cohorts stratified by pharyngeal collapse pattern (AP, lateral, and partial) and BMI category using a BMI threshold of 30 kg/m².

|  |  |  | AP | | Lateral | | Partial | |
| --- | --- | --- | --- | --- | --- | --- | --- | --- |
| Cohort | Parameter | All | High BMI | Low BMI | High BMI | Low BMI | High BMI | Low BMI |
|  | N | 712 | 166 | 316 | 30 | 30 | 70 | 100 |
| Pooled | Percent reduction AHI | 84.7 [62.6, 94.8] | 80.1 [58.1, 91.4] | 87.4 [70.8, 96.4] | 60.2 [35.8, 83.2] | 83.9 [53.9, 90.0] | 80.1 [55.6, 89.2] | 89.0 [71.9, 97.6] |
|  | N Success (%) | 547 (76.8%) | 126 (75.9%) | 251 (79.4%) | 14 (46.7%) | 20 (66.7%) | 52 (74.3%) | 84 (84.0%) |

Table 7. Hypoglossal nerve stimulation outcomes in the pooled cohorts stratified by pharyngeal collapse pattern (AP, lateral, and partial) and BMI category using a BMI threshold of 32 kg/m².

|  |  |  | AP | | Lateral | | Partial | |
| --- | --- | --- | --- | --- | --- | --- | --- | --- |
| Cohort | Parameter | All | High BMI | Low BMI | High BMI | Low BMI | High BMI | Low BMI |
|  | N | 712 | 70 | 412 | 12 | 48 | 31 | 139 |
| Pooled | Percent reduction AHI | 84.7 [62.6, 94.8] | 82.8 [64.7, 91.6] | 85.5 [63.9, 95.9] | 53.9 [34.3, 85.8] | 73.3 [50.5, 89.3] | 78.9 [58.3, 88.7] | 86.2 [66.9, 96.4] |
|  | N Success (%) | 547 (76.8%) | 57 (81.4%) | 320 (77.7%) | 4 (33.3%) | 30 (62.5%) | 23 (74.2%) | 113 (81.3%) |

Table 8. Hypoglossal nerve stimulation outcomes in the pooled cohorts stratified by pharyngeal collapse pattern (AP, lateral, and partial) and BMI category using a BMI threshold of 35 kg/m².

|  |  |  | AP | | Lateral | | Partial | |
| --- | --- | --- | --- | --- | --- | --- | --- | --- |
| Cohort | Parameter | All | High BMI | Low BMI | High BMI | Low BMI | High BMI | Low BMI |
|  | N | 712 | 7 | 475 | 2 | 58 | 3 | 167 |
| Pooled | Percent reduction AHI | 84.7 [62.6, 94.8] | 90.8 [83.7, 97.7] | 85.0 [64.0, 95.3] | 8.8 [-45.8, 63.5] | 72.0 [44.0, 88.9] | 50.0 [-4.5, 85.2] | 85.7 [66.1, 95.7] |
|  | N Success (%) | 547 (76.8%) | 7 (100.0%) | 370 (77.9%) | 0 (0.0%) | 34 (58.6%) | 1 (33.3%) | 135 (80.8%) |
